# Effect of post-activation potentiation on the sports performance of athletes: a systematic review

**DOI:** 10.1101/2023.09.01.23294960

**Authors:** Jiazhe Li, Kim Goek Soh, Su Peng Loh, Shengyao Luo, Marrium Bashir, Xiao Yu

**Affiliations:** Department of Sports Studies, Faculty of Educational Studies, Universiti Putra Malaysia, Serdang, Malaysia; Department of Nutrition, Faculty of Medicine and Health Sciences, Universiti Putra Malaysia, Serdang, Malaysia

**Keywords:** post-activation potentiation, sports performance, athletes, sport, power

## Abstract

**Background:** This study aimed to investigate the effects of post-activation potentiation (PAP) on sport performance.

**Methods:** The data used in this study were reported according to the Preferred Reporting Items for Systematic Reviews and Meta-Analyses 2020 guidelines. Researchers extracted relevant randomized controlled trials from the available literature using prominent scientific indexing databases such as EBSCOhost, PubMed, Scopus, Web of Science, as well as Google Scholar. Ten of the 125 studies that met the inclusion criteria were included. The overall quality of each study was determined using the PEDro scale. Ten studies had scores ranging from 3 to 5.

**Results:** In PAP interventions, power (n = 4) was the primary aspect of sports performance, followed by endurance (n = 1), speed (n = 2), and jumping ability (n = 2). Meanwhile, PAP significantly affects the power, endurance, speed, and jumping abilities of athletes in basketball, volleyball, track and field, and soccer.

**Conclusion:** Compared with conventional training, PAP is a relatively novel dynamic warm-up routine that can greatly enhance the sports performance of athletes in terms of endurance, power, speed, and jumping ability. As a phenomenon of rapid increase in muscle strength and power caused by high-intensity warm-up activities, PAP can help athletes quickly adjust the physical condition to the best state in the pre-match warm-up. However, since some study findings did not reflect the effect of PAP on the sports performance of athletes, more high-quality randomized controlled trials are required to further prove the effect of PAP. Additionally, the existing evidence does not consider the effect of PAP on agility and flexibility performance. In formulating the PAP induction program, it is also necessary to consider the impact of the gender and age of the inducer, exercise level, pre-stimulation load, and interval time on PAP.

**Systematic Review Registration:** [https://inplasy.com/], identifier: [INPLASY202330120].

## 1 Introduction

Tian et al. referred to the abilities of athletes in competitions or training sports performances^1^. This includes the physical fitness, technical and tactical abilities, and mentality of athletes. Optimal physical fitness is the key to improving technical and tactical levels and the requirements of athletes in physical training^2^. Zhang reported that the quality of physical fitness directly determines sports performance, therefore, enhancing physical fitness is essential for enhanced sports performance by athletes^3^.

Physical fitness refers to the essential abilities comprehensively reflected by the functionality of different organs in muscle activity, including five aspects of quality such as speed, strength, endurance, agility, and flexibility^4^. Generally speaking, power can reflect the sports performance of athletes^5–7^, and jumping ability is not only usually uesd to measure athlete’s power^8^, but also the most reliable and valid way to evaluate the power of athlete’s lower limb^9^. Excellent power of athlete’s lower limbs is crucial to the result of a basketball game as it helps the athletes achieve strong jumping ability and acceleration during the game^10^.

There are many ways to enhance sports performance, such as plyometric training, core training and integrative neuromuscular training, etc. PAP is one of the most effective. Post-activation potentiation (PAP) is a phenomenon where voluntary maximal muscular action induces an acute enhancement of previous exercise^11,12^. This activation enhances muscle contraction and potentially increases the performance of solid muscles, particularly in explosion-led sports such as running, jumping, and shooting^13^. The physiological mechanism underlying PAP remains unclear. Nevertheless, previous studies have suggested that PAP has three physiological mechanisms: myosin regulating the enhanced phosphorylation of light chain proteins^14,15^, the nervous system promoting higher-order exercise by increasing unit recruitment, and alterations in muscle fiber pinnate angles^11,16,17^.

The results show that improved phosphorylation of light chain proteins after maximal (and even submaximal) stimulation results in improved muscle contraction ability ^18,19^. This mechanism regulating light chain phosphorylation can make myosin-actin interactions more sensitive to calcium ions. The myosin light-chain-activating enzyme produces more adenosine triphosphate in the myosin complex. The frequency of myosin cross-bridge oscillations then increases; thus, pre-stimulation increases the cross-bridge power output, which improves exercise performance^14^.

Another possible mechanism for the development of PAP is the increased involvement of the nervous system in skeletal muscle activity, which increases the degree of synchronous motor fiber contraction and the number of higher-order motor units involved in the movement^16,20^. Enhanced neural activity may result from increased motor-unit recruitment, improved motor-unit synchronization, reduced synaptic inhibition, or increased central nerve impulse inputs^21,22^. Fewer motor neurons control slow-twitch muscle fibers during pre-stimulation and low-load stimulation exercises. As the pre-stimulus load increases, higher-order motor units (fast muscles) controlled by large motor neurons are recruited.

Tillin and Bishop suggested that recruiting many higher-order motor units and acting on the nervous system may be one of the mechanisms of post-activation enhancement^11^.

However, changes in the pinnate angle of the muscle fibers may also contribute to the promotion of PAP. Folland and Williams showed that the pinnate angle affects the force transmitted by the muscle to the tendons and bones^23^. Because a smaller pinnate angle decreases force transmission, muscle contraction torque increases as the pinnate angle decreases. Mahlfeld et al. reported that static maximal voluntary contraction, including stimulation, can promote pinnate angle reduction^24^. Kumagai et al. suggested that narrowing the feature angle could enhance sprint speeds in athletes^25^. However, a previous study showed that a larger feature angle can improve power performance^26^.

Benefits of PAP stimulation, such as enhancing sprint performance, jumping ability, and other strength-related movements, have been reported by several studies^27–34^. Therefore, for a particular individual, the optimal duration and intensity of PAP stimulation should be considered, combined with subsequent rest periods, because fatigue may outweigh the potential benefits if not properly prescribed.

PAP is affected by numerous factors during the induction process, such as the pre- stimulation approach and load, interval time, participant characteristics, and evaluation techniques^35–38^. Currently, there are many studies on PAP and its influencing factors, but some studies remain controversial regarding whether PAP can effectively improve the sports performances of athletes. Additionally, there is still a need for a clear conclusion on how to effectively induce and maximize the effects of PAP. Liang et al. suggested that accurate control of the interval time is the key to achieving the best effect^39^. The window time occurs when PAP is greater than the fatigue effect. Motor performance improvement caused by PAP appears during two window-opening periods during induction exercises. At the beginning of the induction exercise, there is usually a window-opening period (first); however, the enhancement effect of PAP is not apparent due to the current small amount of exercise and intensity, and the duration of PAP is relatively short. As the exercise load increases, the fatigue effect gradually increases, which can reduce sports performance when explosive activity is performed. A few minutes after the end of the induction exercise, as the fatigue effect gradually subsides, PAP becomes more significant than the fatigue effect. The second window-opening period is when sports performance improves most significantly.

As shown in Fig 1, the enhancement effect of PAP in the second window-opening period is more evident than that in the first, and the second window also lasts longer than the first window^11^. Tillin and Bishop suggested that PAP was the “potential difference” between the enhancement and fatigue effects after the induced exercise stimulated the muscle^11^. Only when the PAP is higher than the fatigue effect will the muscle contraction traces significantly improve the subsequent performances of athletes. The timing, duration, and intensity of PAP depend on the balance between enhancement and fatigue effects, which affects a series of factors. Therefore, this study adopted a systematic review method to analyze experimental research reports on the impact of PAP on the sports performance of professional or amateur athletes. This study also aims to test the exact effect of PAP on the different performance indicators of the athletes’ sports performance and provide a reference for the training practices of athletes in different sports.

**Fig 1.**
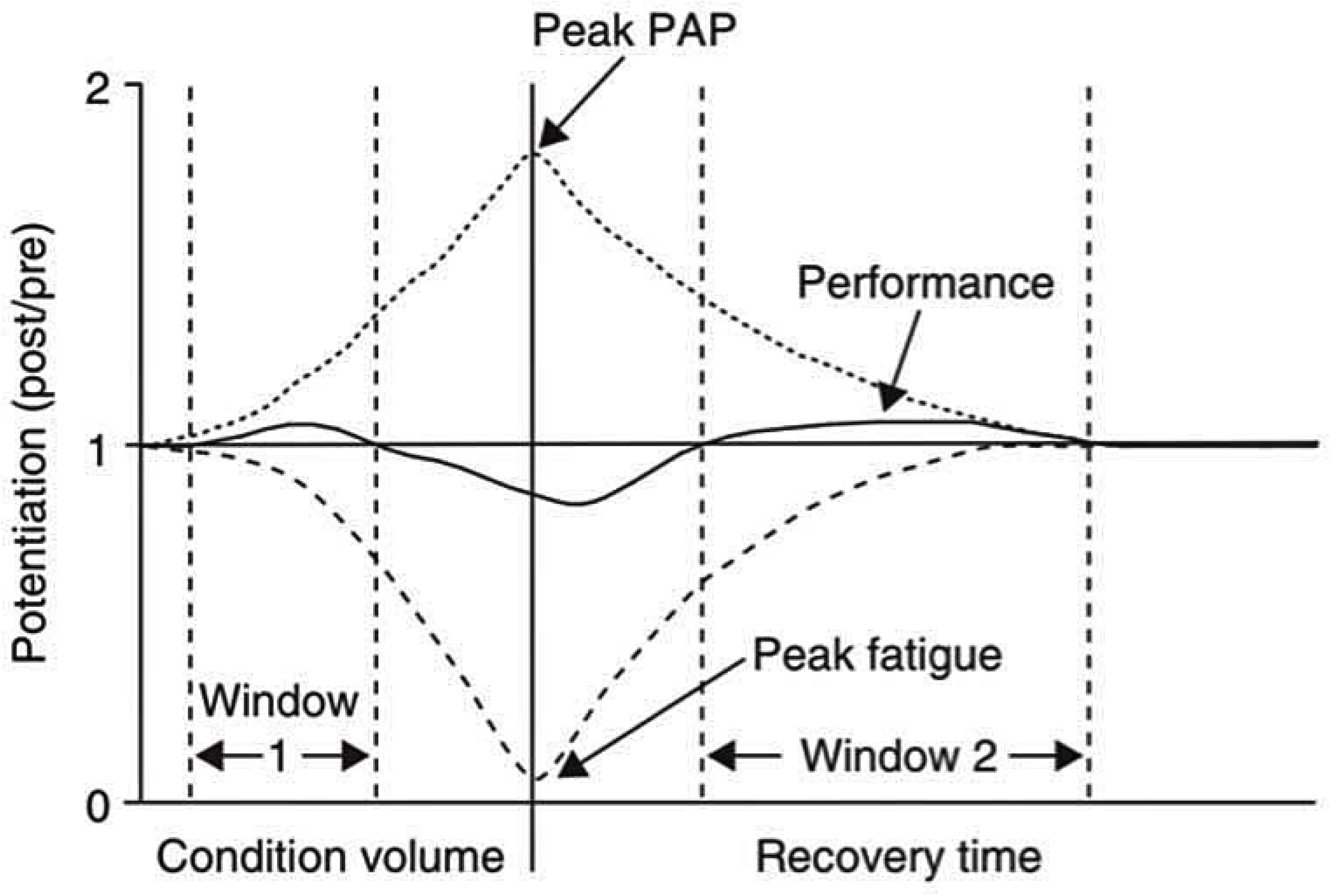
Relationship of post-activation potentiation with fatigue^11^. Descriptive text : Fig1 provides a model that introduces the hypothesized relationship between PAP and fatigue, with PAP dominating more than fatigue when the pre-activation(condition) intensity is low, followed by a rapid increase in muscle movement during subsequent reinforcemen (post/pre) (window 1). As the pre-activation intensity increases, fatigue becomes dominant, negatively affecting subsequent sports performance. Following the pre-activation, fatigue dissipates at a faster rate than PAP, and a potentiation of subsequent enhanced performance can be realized at some point during the recovery phase (window 2).

## 2 Materials and Methods

### 2.1 Protocol and Registration

This study used the Preferred Reporting Items for Systematic Reviews and Meta-Analyses (PRISMA) framework to gather and assess data. It was registered in INPLASY (registration number: INPLASY202330120, DOI number: 10.37766/inplasy 2023.3. 0120)^40^.

### 2.2 Search Strategy

Popular scientific databases were used to search for relevant literature, including Ebscohost, PubMed, Scopus, Web of Sciences, as well as Google Scholar, up to October 10, 2022. For each independent search engine, the title and abstract were used as strategic search queries. The main keywords used for collecting relevant research works were: (“PAP” OR “Post-activation potentiation” OR “Pre-activation” OR “Pre- conditioning”) AND (“Sports Performance” OR “Athletic Performance” OR “Power” OR “Endurance” OR “Jumping Ability” OR “Agility” OR “Speed” OR “Flexibility”) AND (“Athlete*” OR “Player*” OR “Sportsman*” OR “Sportswoman*” OR “Sportsperson*” OR “Jock*”).

### 2.3 Eligibility Criteria

The population (P), intervention (I), comparison (C), outcome (O), and study design (S) (PICOS) model was used in the literature. Each PICOS factor was used as an inclusion criterion for the retrieved publications. We included studies that met the following inclusion criteria: (P) The study population consisted of professionally trained and untrained healthy personnel, regardless of gender or age; (I) PAP should be isolated and expressly discussed, with PAP in the experimental group and other training methods compared to no training in the control group; (C) The comparisons in this study involved both single- and multiple-group trials; (O) The findings of this study revealed at least one post-activation performance enhancement effect in athletes; and (S) All experimental designs included in the literature were randomized controlled trials.

### 2.4 Study Selection

Two authors individually selected the studies that met the inclusion criteria. The abstracts and titles of studies were assessed to identify those that could be incorporated into the study. In response to a debate between the two authors regarding the selection of a paper, an additional author was requested to assess the entire work before arriving at a definitive conclusion.

### 2.5 Data Extraction and Quality Assessment

After the screening, we acquired the following crucial data from several eligible studies: (1) author name and publication year; (2) demographic characteristics such as the number, type, gender, and age of the participants; (3) intervention characteristics such as the type and measurement indicators, interval time, and duration; and (4) study results.

The PEDro scale is a valid indicator of the methodological quality of systematic reviews owing to its high validity and reliability^41^. There were 11 items on the scale, with scores ranging from 0 to 10. Two independent raters scored the 11 items as “yes” (1 point) or “no” (0 point). A third rater addressed any discrepancies in the rating procedure.

However, eligibility criteria were not factored into the overall external validity scores. The total PEDro score ranged from 0 to 10. The higher the grade, the higher the quality of the method. The quality of the method is proportional to its PEDro score. Studies rated 8 to 10 were regarded as methodologically superior. Studies rated 5 to 7 were of high quality, those rated 3 to 4 were of average quality, and those ranked below 3 were of poor quality^42^. Overall, the scientific evidence was evaluated based on three levels: quantity, methodological quality, and consistency of research findings.

Generally, substantial evidence provides consistent information regarding the number and results of multiple studies. Moderate evidence when a single study exists or when information from multiple studies is inconsistent. There is no evidence when there are no case-control studies available.

## 3 Results

### 3.1 Study Selection

Fig 2 illustrates the literature filtering technique. After the initial evaluation, 120 studies were retrieved. This number was determined after Endnote software deleted duplicate studies, followed by a second round of removal that included three non-full-text papers, two non-English studies, and nine non-journal studies. In the third screening phase, 106 full-text studies were evaluated for eligibility. Of these, 89 were excluded because they did not satisfy the inclusion criteria for the randomized controlled trial; four were excluded because they were irrelevant to the topic; and three were discarded. The qualitative analysis examined ten relevant publications that met the inclusion criteria.

**Fig 2.**
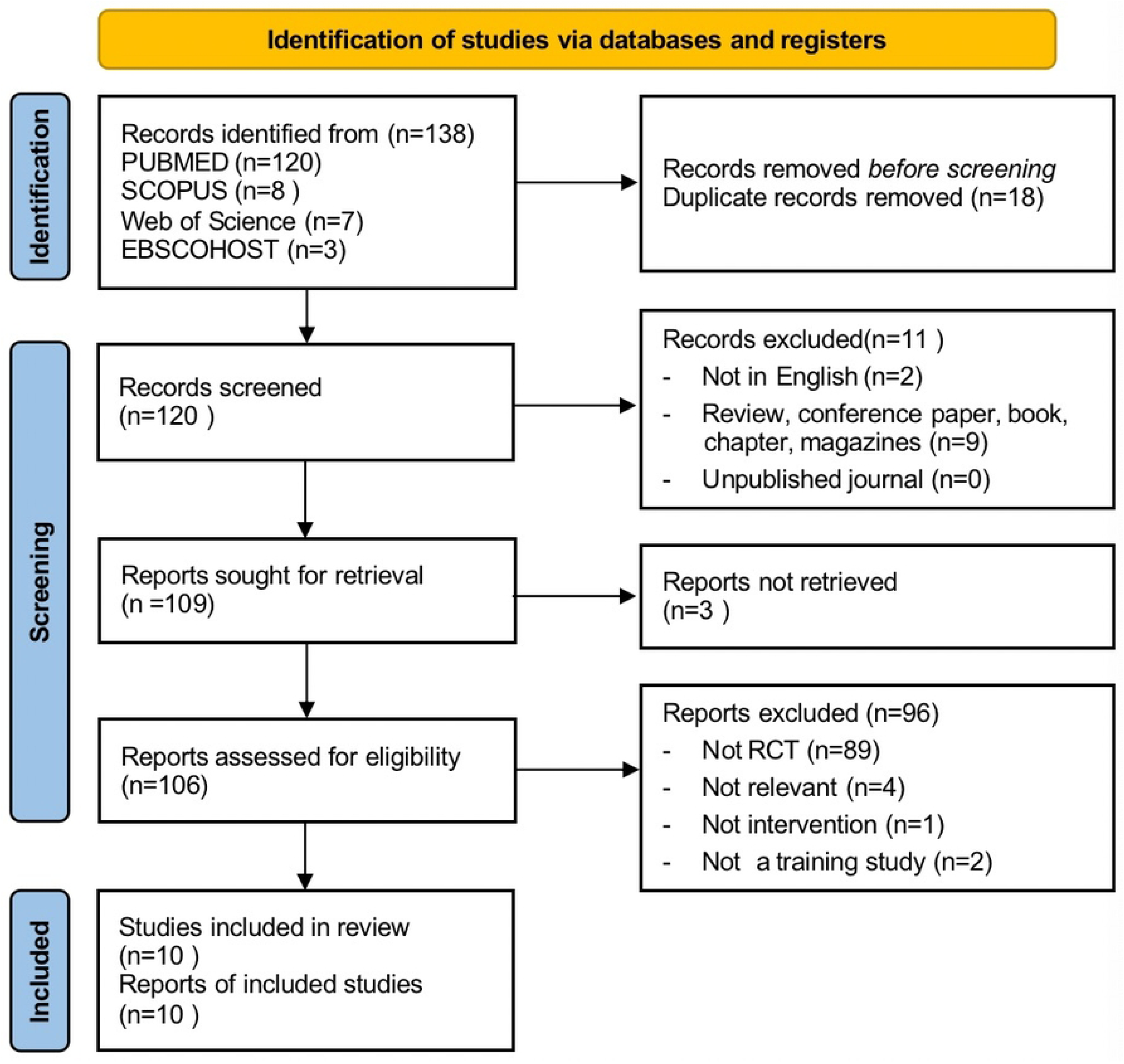
The search, screening and selection processes for suitable studies–based on PRISMA^43^. Descriptive text:Fig 2 depicts the literature review process.

### 3.2 Study Quality Assessment

The PEDro scale scores for each study are presented in Table 2. All study data were evaluated on a 3- to 5-point PEDro scale, penalizing for assignment concealment, blinding the evaluator, participants, therapists, and intent-to-analyze criteria. The intervention in this study involved resistance training, which can be challenging for blind participants, evaluators, and clinicians and carries the risk of sports injuries and professional misconduct. However, our study ensured that all participants were treated equally. Two studies with scores of less than four were disqualified^44,45^.

**TABLE 1.** PICOS Eligibility criteria.

| PICOS | Detailed Information |
| --- | --- |
| Population | Healthy players |
| Intervention | Post-activation potentiation |
| Comparison | Single-group trials, Two groups, and Three groups |
| Outcome | Include post-activation potentiation with various kinds of sports performance among players |
| Study Design | Single-group Trials or Randomized Controlled Trials |

**TABLE 2.** Summary of methodological quality assessment scores.

| Study | Eligibility<br>Criteria | Random<br>Allocation | Allocation<br>Concealment | Baseline<br>Comparability | Blind<br>Participants | Blind<br>Therapist | Blind<br>Assessor | Follow-Up | Intention to<br>Treat<br>Analysis | Between<br>Group<br>Comparisons | Point<br>Measure and<br>Variability | Total PEDro<br>Score |
| --- | --- | --- | --- | --- | --- | --- | --- | --- | --- | --- | --- | --- |
| Marco et al. (2019) | 0 | 0 | 0 | 0 | 0 | 0 | 0 | 1 | 0 | 1 | 1 | 3 |
| Lamberto et al. (2022) | 1 | 1 | 0 | 1 | 0 | 0 | 0 | 1 | 0 | 1 | 1 | 5 |
| Jonathan et al. (2019) | 0 | 0 | 0 | 1 | 0 | 0 | 0 | 1 | 0 | 1 | 1 | 4 |
| Danny et al. (2017) | 0 | 0 | 0 | 0 | 0 | 0 | 0 | 1 | 0 | 1 | 1 | 3 |
| Mariola et al. (2020) | 0 | 0 | 0 | 1 | 0 | 0 | 0 | 1 | 0 | 1 | 1 | 4 |
| Joseph et al. (2010) | 0 | 0 | 0 | 1 | 0 | 0 | 0 | 1 | 0 | 1 | 1 | 4 |
| Matthew et al. (2010) | 1 | 1 | 0 | 1 | 0 | 0 | 0 | 1 | 0 | 1 | 1 | 5 |
| Jameson et al. (2014) | 1 | 1 | 0 | 1 | 0 | 0 | 0 | 1 | 0 | 1 | 1 | 5 |
| Mauro et al. (2018) | 1 | 0 | 0 | 1 | 0 | 0 | 0 | 1 | 0 | 1 | 1 | 4 |
| Ceith et al. (2017) | 1 | 1 | 0 | 0 | 0 | 0 | 0 | 1 | 0 | 1 | 1 | 4 |

### 3.3 Participant Characteristics

Table 3 provides the details of the participants in the studies that met the inclusion criteria.

1. Classification of athletes. Among the 8 studies, two were on soccer players^46,47^; two on volleyball players^48,49^; one on basketball players^50^; and the remaining five on rugby^51^, track and field^52^ and endurance-trained athletes^53^.
2. Gender, Number, and Age. The study included a total of 109 participants, including 52 males and eight females, with gender details not provided for the remaining 49 participants^46,48,50^. The studies reported the age range of the participants as being between 16 and 29 years old.

**TABLE 3.**
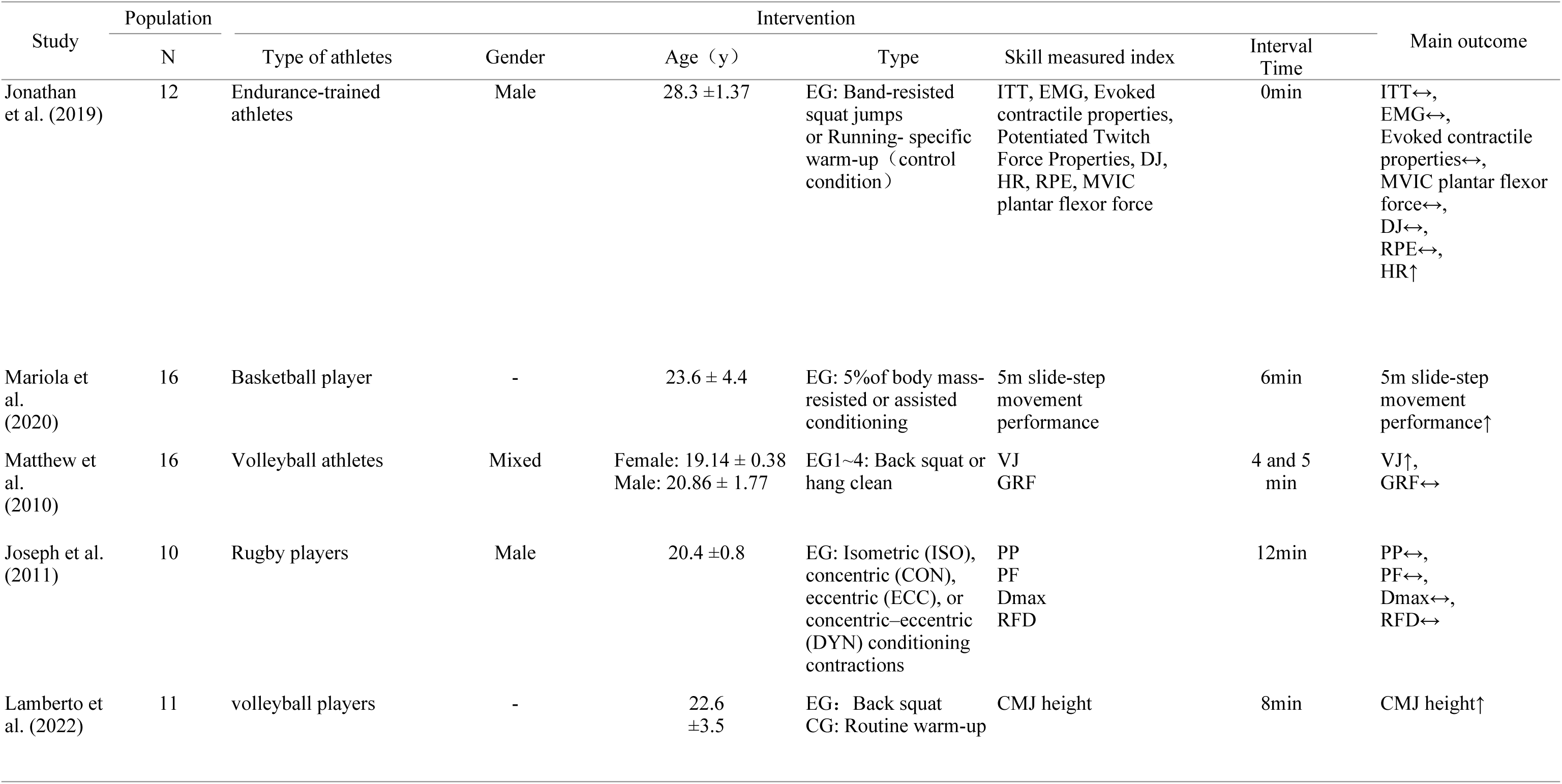
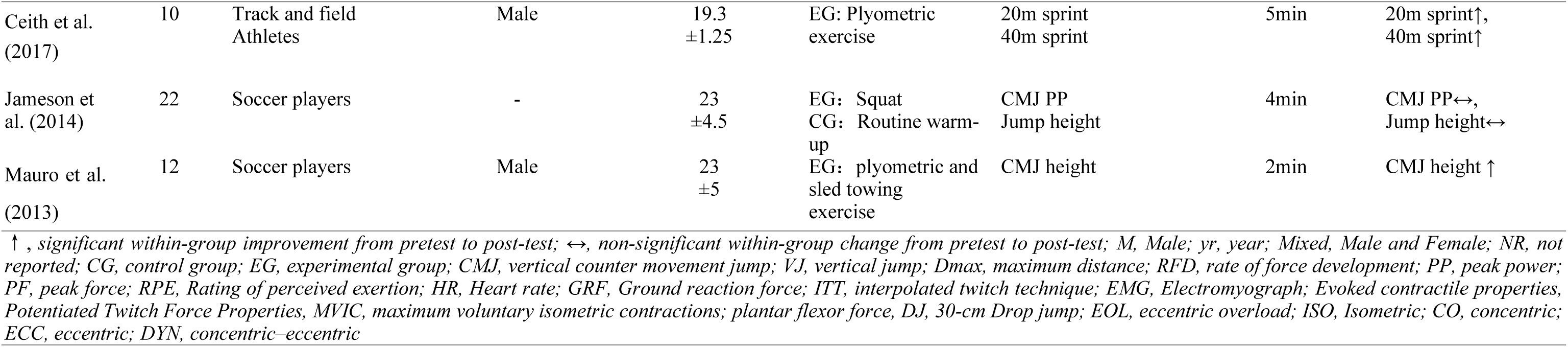
Population, Intervention and Main outcome.

### 3.4 Intervention Characteristics

The eight included intervention characteristics were reported based on several properties:

1. Mode of pre-stimulus. Three studies selected back squats^46,48,49^; one study used a squatted jump^53^; and another study used a bench press as pre-stimulus modes^51^. The plate jump method was used for pre-stimulation^52^. The pre-stimulus mode chosen by Mauro et al. was more complicated, combining plyometric exercises and sled towing, whereas Mariola et al. adopted a consistent pre-stimulus mode with the test method^47,50^.
2. Pre-stimulus intensity. Lamberto et al. chose a high-intensity load of three sets of 90% repetition maximum (RM) (1RM), and two studies chose a relevant load intensity: four sets of 5RM, three sets of 3RM, and one set of 3RM^48,51,53^. Matthews et al. reported a mixture of moderate- and maximum-intensity stimulations consisting of five sets of 50% 5RM and five sets of 80% 5RM back squats^49^. Additionally, two studies reported stimulation with lower loads: one set of 3RM and a mass of 11.2 kilograms, respectively^46,52^. Another study reported that the pre-stimulation modality is relatively complicated. The conditioning stimulus comprised two sets of 15 ankle hops, three sets of five hurdle hops, and three sets of 20-meter sprints and sled towing, amounting to 45 jumps and 60 meters of sprinting with an external load^47^.
3. Interval time. These studies tested the measures every 2 minutes, 4 minutes, 4 and 5 minutes and 5 minutes, 6 minute, 8 minutes, 12 minutes after pre-stimulation. Notably, only one study reported no rest interval (0 minute)^46–53^.

### 3.5 Outcome

The results of this study were grouped according to the impact of PAP on athletic performance. The authors of this study independently categorized studies based on other research areas. The staging was passed, and discussions between the authors were resolved until a consensus was reached through consultation.

#### 3.5.1 Effect of PAP on Power

Among the eight studies included in this review, six reported^46–49,51,53^. The aspects valued and assessment tools used were the reactive strength index test, power performance test (peak power [P_peak_], maximum distance [D_max_], peak force [F_peak_], and rate of force development [RFD]), and countermovement jump test (jump height, ground reaction force [GRF], and P_peak_)^46,49,51,53^. The participants included 12 healthy endurance-trained athletes, ten male competitive rugby athletes, 16 mixed-gender volleyball athletes, 11national Superliga-2 volleyball players, 12 professional male soccer players, and 22 senior professional soccer players^46–49,51,53^.

On the one hand, two studies examined the effect of post-activation potentiation on power among 27 volleyball athletes^48,49^. The result of the first group investigated the condition that produced the greatest increase in vertical jump height, which resulted in an average increase of 5.7% (2.72 ± 1.21 cm; p <0.001), but there was no significant difference in peak GRF (p >0.05)^49^. Meanwhile, another study revealed that there was a significant improvement among the control group and experimental group from PAP (35.40 ± 3.69 vs. 29.61 ± 4.10 cm; p <0.05), pre-Match (37.10 ± 4.09 vs. 31.38 ± 3.99 cm; p <0.05), Set 1 (38.84 ± 4.74 vs. 31.22 ± 2.61 cm; p <0.05), Set 2 (41.37 ± 4.91 vs. 32.75 ± 4.47 cm; p <0.05). Additionally, a significant improvement was observed in the countermovement jump test in the experimental group between baseline (pre-PAPE) and all the following tests: post-PAPE (+1.3 cm), pre-match (+3.0 cm), Set 1 (+4.8 cm), Set 2 (+7.3 cm), Set 3 (+5.1 cm), Set 4 (+3.6 cm), and Set 5 (+4.0 cm). The jump height in the control group was lower than that in the experimental group^48^.

On the other hand, two studies were related to soccer power performance, involving 34 professional soccer players^46,47^. The first group of studies investigated the impact of PAP on power by using a countermovement jump test on 12 professional male soccer players^47^. The authors concluded that PAP could significantly improve countermovement jump height in soccer athletes at T1, T3, and T5 (expressed in minutes) under caffeine conditions (5.07%, 5.75%, and 5.40% increase, respectively; p <0.01) compared to baseline.

In addition, one study found no statistically significant differences in the countermovement leap test (P_peak_, p >0.05 and jump height, p >0.05)^46^. Meanwhile, in a different study, PAP produced by isometric (ISO) training was substantially greater (587 <u>±</u> 116 and 605 <u>±</u> 126 W for pre- and post-ballistic bench press throw, respectively; p >0.05). And there was no significant differences in P_peak_ for concentric (CON), eccentric(ECC), and concentric-eccentric(DYN)(p >0.05), and there were no significant differences existed in F_peak_, D_max_, and RFD (p >0.05) for ISO, CON, ECC, and DYN. And no significant improvement in EMG were found between pre- and post-BBPT for all of the pre-activation mode^51^. In addition, One study revealed that PAP had a positive effect on the reactive strength index (p = 0.035; 3.6%) compared with control conditioning^53^.

#### 3.5.2 Effect of PAP on Speed

Speed was considered in two of the eight studies included in this review^50,52^. The measurement tools were a 5-meter slide-step movement test and 20- and 40-meter sprint tests. The participants included 16 basketball players and ten male track and field athletes ^50,52^. One study investigated two exercise protocols, namely, assisted and resisted conditioning ability (CA), each comprising four sets of 10-meter slide-step movement with a body mass external load of 5% and one-minute rest intervals. The result indicated a statistically significant difference between baseline and post-assisted post-activation performance enhancement for the experiment group in the 5-meter slide- step movement time (3.09 ± 0.16 vs. 3.24 ± 0.15 s; p <0.05). However, the other exercise protocol reported no significant difference between baseline and post-intervention after the resistance CA (3.23 ± 0.15 vs. 3.17 ± 0.13 s; p = 0.230)^50^. Additionally, Ceith et al. observed a decrease in sprint time when PAP in the form of a plyometric was performed during warm-up for both 20-meter sprints (3.134 vs. 3.172 s; p <0.05) and 40-meter sprints (5.337 vs. 5.405 s; p <0.01)^52^.

#### 3.5.3 Effect of PAP on Endurance

Only one study included in this systematic review investigated the relationship between PAP and endurance performance^53^. The time it took 12 endurance-trained male athletes (28.33 ± 1.37 years) to complete the time-trial test determined their endurance. Jonathan et al. reported that PAP stimulus decreased the time required to run by 3.6% (p = 0.07) and 8% (p = 0.014) over a distance of one kilometer^53^.

## 4 Discussion

### 4.1 Effect of PAP on Power

Power determines sports performance^54^. Meanwhile, in studies of power performance, jumping performance is a measure method that frequently is used^55^. Studies have shown that PAP can be generated through induction training, thereby improving performance in explosive sports. Based on the six studies that analyzed the effectiveness of PAP in improving the power performance of athletes, we draw firm conclusions. Three studies indicated that PAP can improve the power performance of athletes, which is consistent with the findings of Gourgoulis et al. and Webber et al.^47,48,53,56,57^. However, further studies are required to support the idea that PAP can improve the power performance of athletes.

Joseph et al. proposed several power evaluation methods, such as P_peak_, F_peak_, D_max_, and RFD^51^. Several different PAP activation methods such as ISO, CON, ECC, or DYN conditioning contractions were used, in which the effects of PAP produced by ISO conditioning contractions showed a significant increase in P_peak_, but no significant differences in P_peak_ were found for CON, ECC, or DYN conditioning contractions. F_peak_, D_max_, and RFD did not differ significantly among the ISO, CON, ECC, and DYN groups. This result may be primarily attributable to the time interval and pre-stimulus interaction of PAP^58,59^. Matthews et al. found a significant increase in vertical leap height but no difference in peak GRF^49^. This result may be due to the various training experiences and pre-stimulation load intensities of the participants^60,61^.

In addition, Jameson et al. reported that no effect was observed for P_peak_ or jump height during experimental group trials^46^. Baechle and Earle reported that this was likely due to the determination by 3RM of the participants and testing within the same day, which resulted in fatigue of the central nervous system^62^. At the same time, this result is similar to that of Khamoui et al., who suggested that PAP did not significantly improve sports performance^63^. The reason for this may be that when the activation method is inconsistent with the specific movement of the athlete, it destroys the neuromuscular contraction memory of the particular exercise in the previous session, which adversely affects subsequent sports performance and fails to activate PAP^63^.

### 4.2 Effect of PAP on Speed

Of the eight studies included in this review, two involved speed tests, and both consistently suggest that PAP can improve the speed performance of athletes^50,52^. Studies have also shown that PAP can improve rapid skeletal muscle strength, and increased neural activation of motor units during the potentiation phase may be the primary reason for the enhanced speed performance, which is consistent with the findings of Healy and Comyns and Lesinski et al.^64,65^. Notably, Mariola et al. reported a new finding that only 5% of body mass-assisted CA significantly enhanced the overall performance of the slide-step movement^50^. However, some previous studies, including Comyns et al., Crewther et al., and McBride et al., concluded that lower stimulus loads were not effective in increasing sprint speed^66–68^. In contrast, some other studies have reported that light-resistance-assisted conditioning can effectively elicit potentiation^69–71^.

In response to this change, Ceith et al. suggested that low-intensity pre-stimulation would produce sufficient central nervous system stimulation without leading to a high degree of fatigue^52^. PAP is a personalized and complex phenomenon. Although two studies have shown that PAP can improve speed performance in athletes, due to the absence of physiological analysis, the cause of these changes has yet to be identified or explained. PAP can be affected by the gender, age, muscle fiber type, and training experience of the participants^72–74^. Moreover, the electromyograph of stimulated muscles has not been studied with kinematic and kinetic data analysis. The assessment of performance changes based on a single resistive value makes it difficult to draw definitive conclusions. Therefore, when formulating a PAP plan, individual differences among athletes and the diversity of various movements should be considered. The activation method and load intensity should be arranged according to the characteristics of the athletes and special projects to minimize the factors that may negatively affect PAP.

### 4.3 Effect of PAP on Endurance

According to a study by Jonathan et al., PAP may help endurance athletes improve their endurance performance and neuromuscular properties during long-distance running, which is supported by Silva et al. and Feros et al.^53,75,76^. Both studies confirmed that PAP significantly affected endurance performance. Several studies have shown that the skeletal muscles can adjust peripherally to reduce fatigue and improve exercise performance^77,77,78^. This adjustment may explain why PAP improves endurance performance in athletes.

It should be noted that the participants in this study were endurance-trained athletes, so the difference in PAP or fatigue effect among muscle phenotypes can be more pronounced since only slow-twitch fibers can maintain Ca^2+^ sensitivity after prolonged endurance activity and after appropriate CA^79^. The more potent antifatigue properties of long-term skeletal muscle training may produce a more intense strengthening effect.

Therefore, the included studies only reported the effects of PAP on athletes with endurance training experience, which is an essential gap in the literature. Athletes with different characteristics have different effects due to the influence of PAP. Therefore, it is necessary to consider individual differences among athletes in research on the effects of PAP on endurance performance and to develop specific PAP programs.

## 5 Limitation

Although this study offers evidence to assess the effects of PAP on sports performance, some of the weaknesses revealed by its limitations are as follows: First, according to the currently retrieved studies, this work on the effect of PAP on the sports performance of athletes can only be limited to the effect of PAP on their physical fitness and does not involve studies on their skill performance or performance throughout the game. Second, the training level of athletes is often evaluated as a moderating factor affecting the effect of PAP. The data from PAP studies in the existing literature is all based on the training experience of the athletes to reflect their training level, never considering aerobic or anaerobic capacity. Jones et al. suggested that recovery from fatigue after pre-stimulation is an aerobic phenomenon, creating a gap in existing research^80^. Third, studies have demonstrated that PAP stimulation leads to fatigue. Skeletal muscle fatigue and strength enhancement coexisted during PAP induction. A short interval produces a good effect of PAP only if the degree of fatigue is within the enhancement effect^81,82^.

Fourth, the content of each training objective in the included literature varied, and the evaluation methods were diverse, making it impossible to conduct an in-depth analysis of the intensity and quantity of the intervention. Fifth, as the existing literature is based on athletes with training experience, the results of these studies cannot provide a reference for untrained participants. The existing literature is based on male athletes, and there is only one study with mixed genders; therefore, the results of these studies lack research on the impact of female athletes on sports performance. In addition, existing literature does not consider the effect of PAP on agility and flexibility performance. Whether PAP improves the agility and flexibility of athletes requires further research.

## 6 Conclusion

This review reported that PAP can be effectively induced by either high-intensity or medium- and low-intensity induction, and rest intervals between 2 to 8 minutes can significantly improve the sports performance of the subjects.

In addition, existing studies provides evidences that PAP can improve athletic power, speed and endurance performance. Traditional warm-up methods such as jogging and stretching have been proven to have limited effects on improving athletes’ sports performance. Adding PAP to the pre-match warm-up can enhance the muscle working ability of athletes in a specific time period and improve sports performance. However, PAP improves sports performance, the outcomes could be a result of factors like gender and age, type of muscle fiber, training status, and the training level of the athletes. At the same time, the optimal interval time for PAP has yet to be determined. Therefore, different intervals lead to different conclusions in the existing literatures.

When formulating an induction PAP program, it is crucial to investigate the influence of individual differences in participants on the effect of PAP. Therefore, researchers must continue to explore these gaps to develop more scientific training plans. This study recommends that trainers and researchers consider the effects of the motor characteristics of the individuals and their differences in PAP when designing pre-stimulation protocols for PAP.

## 8 Author Contributions

JL drafted the article. JL and KS provided data interpretation. KS critically revised the article and gave the final approval. All the authors read and approved the finial manuscript.

## 10 Data Availability Statement

The original contributions presented in the study are included in the article/Supplementary Material, further inquiries can be directed to the corresponding authors.

